# Passive Advancement of the Aspiration Catheter During Mechanical Thrombectomy is Independently Associated with Superior Recanalization Outcomes

**DOI:** 10.64898/2026.09.20.26363528

**Authors:** MD Alexander, RS Khangura, B Varjavand, TA Chaudhry, WT Kim, F Settecase

**Affiliations:** Sutter Medical Center Sacramento, UNITED STATES; Sutter Health Valley Area, UNITED STATES; California Pacific Medical Center

**Keywords:** passive advancement, mechanical thrombectomy, ASCENT, aspiration catheter, first-pass effect, recanalization, secondary occlusion, Tenzing

## Abstract

**Background:** Passive advancement—antegrade movement of the aspiration catheter upon inner delivery device withdrawal without active operator force—is a physical phenomenon observed during mechanical thrombectomy, with anecdotal reports of favorable recanalization outcomes. This study characterizes its prevalence and associations with recanalization.

**Methods:** Consecutive thrombectomy cases at two high-volume stroke centers (January 2019– December 2025) were retrospectively reviewed. Stored fluoroscopy and angiograms were assessed for passive advancement and final catheter tip position at arterial branch points. Per-pass and per-vessel mixed-effects multivariable regression identified independent associations with eTICI score, first-pass effect (FPE, eTICI ≥2c-3), modified FPE (mFPE, eTICI ≥2b-3), secondary occlusion, and complications.

**Results:** Of 1,824 passes in 995 consecutive patients (1,027 occlusions), 733 had imaging adequate for passive advancement assessment. Passive advancement occurred in 525/733 passes (71.6%), including to an arterial branch in 306/733 (41.7%). On per-pass analysis, passive advancement was associated with higher eTICI ≥2b-3 (71.0% vs 19.7%) and eTICI ≥2c-3 (57.5% vs 10.1%); advancement to a branch amplified these associations (90.5% vs 32.1% and 79.4% vs 18.7%; all p<0.001). Among 429 first passes, mFPE was 74.1% vs 10.8% and FPE 63.4% vs 7.5% with vs without passive advancement; rates reached 95.0% and 87.2%, respectively, at a branch point (all p<0.001). Passive advancement to a branch was inversely associated with secondary occlusion (13.3% vs 31.3%; p<0.001). On multivariable analysis, passive advancement independently predicted eTICI score, eTICI ≥2b-3, and eTICI ≥2c-3 (all p<0.001); final catheter position at a branch was the dominant independent predictor of FPE, mFPE, secondary occlusion, any complication, and intracranial complication on per-vessel analysis (all p≤0.046).

**Conclusions:** Passive advancement is a prevalent physical phenomenon and an independent predictor of successful recanalization. When advancing to an arterial branch point, it is associated with mFPE >87%. Passive advancement should be a technical objective of aspiration thrombectomy.

**KEY MESSAGES:** *What is already known on this topic:* Catheter tip position relative to the thrombus and target vessel anatomy affects recanalization success in aspiration thrombectomy. Maximizing outer catheter-to-vessel inner diameter ratio and achieving distal catheter positioning improve aspiration success. Secondary occlusion is an important complication and can be caused by clot fragmentation during thrombectomy.

*What this study adds:* This manuscript describes and systematically characterizes a physical phenomenon— passive advancement of the aspiration catheter upon inner delivery device withdrawal— that occurs in 71.6% of assessed thrombectomy passes and is independently associated with higher rates of successful recanalization. When passive advancement carries the catheter to an arterial branch point, first-pass complete reperfusion (eTICI ≥2c-3) is achieved in 87.2% of cases. Final catheter position at an arterial branch is the dominant independent predictor of recanalization success, secondary occlusion prevention, and complication avoidance on per-vessel multivariable analysis.

*How this study might affect research, practice or policy:* Operators performing aspiration thrombectomy should recognize passive advancement of the aspiration catheter as a favorable prognostic sign and allow this to occur during inner device withdrawal, including allowing the catheter tip to passively advance to an arterial branch point. Future thrombectomy device designs, operator training protocols, and procedural guidelines should include passive advancement as a primary technical objective, and prospective studies should validate these observations.

## INTRODUCTION

Mechanical thrombectomy techniques have changed rapidly since the release of the first positive randomized trials results in 2014.^1–2^ Contemporary aspiration-based techniques, including ADAPT (A Direct Aspiration First Pass Technique) and ASCENT (ASpiration Clot Extraction Navigated by Tenzing), have achieved recanalization rates that exceed those reported in early landmark stent retriever (SR) trials.^3–5^ A key determinant of this progress has been recognition that outer catheter-to-vessel inner diameter matching is a primary mechanical driver of aspiration success. Maximizing the catheter-to-vessel ratio achieves near-complete distal flow arrest on catheter insertion, optimizing clot extraction forces and minimizing fragmentation and distal embolization.^6–7^

A new generation of inner delivery devices such as the Tenzing device (Route 92 Medical, West Jordan, UT) reduce the ledge effect caused by step-off between the aspiration catheter and its inner delivery device, enabling more effective navigation of large-bore and super large-bore aspiration catheters to the clot face while avoiding the crossing of the thrombus with inner devices.^8–9^ Operators experienced with this system have anecdotally noted a physical phenomenon in which the aspiration catheter moves antegrade within the target vessel upon inner delivery device withdrawal, without actively applied operator force on the aspiration catheter, an aspiration catheter behavior we herein term passive advancement. Advancement of a catheter over an inner microcatheter is a common occurrence in endovascular procedures. As the innermost device is withdrawn, device removal can reduce accumulated friction and cause straightening in redundant curves along the course of the devices. With a fixed length of outer catheter now traversing across a shorter distance, this can cause the outer catheter distal tip to move antegrade within the vessel. In general, neuroendovascular operators are trained to recognize this phenomenon and mitigate or counteract it, to prevent catheter movement that may not be desirable or to prevent potential vascular injury. Passive advancement appears to occur more readily as a tapered inner delivery device is removed from an aspiration catheter during mechanical thrombectomy. This likely relates to the lubricious coating of these delivery devices and the reduced annular space between the outer margin of the inner device and the inner wall of the aspiration catheter. Moreover, experienced operators have anecdotally noted a particularly strong association between catheter passive advancement to an arterial branch point and favorable recanalization. It has been hypothesized that a vacuum generated during removal of a space-occupying inner delivery device from an aspiration catheter may cause initial clot ingestion into the aspiration catheter prior to application of pump or syringe vacuum suction, increasing the probability of recanalization.^10^ Passive advancement and the vacuum created during delivery device removal appear to work synergistically to potentiate clot removal.

Mechanistic support for this phenomenon exists. Bench testing of the Tenzing delivery device withdrawal in a physiologic three-dimensional circle of Willis model documented a transient negative pressure spike—reaching 52–97% of maximum suction vacuum—at the clot-catheter interface during inner device removal that produced clot mobilization before active aspiration commenced.^10^ This pre-aspiration clot engagement or synthetic aperture aspiration effect has been proposed as the mechanism by which the inner device primes aspiration efficiency. To date, however, the clinical prevalence of passive advancement as a directly observable fluoroscopic event, its degree (partial passive advancement vs passive advancement to an arterial branch), and its independent associations with recanalization have not been previously characterized.

Catheter tip position relative to target vessel anatomy has been previously shown to matter. Distal access catheter tip positioning is associated with superior reperfusion in combined stent retriever-aspiration approaches.^11^ The degree to which advancement to an arterial branch specifically modifies outcomes has not been examined.

This study determines the prevalence passive advancement across a consecutive thrombectomy series, and assesses its association with recanalization, secondary occlusion, and procedural safety on per-pass and per-vessel basis.

## METHODS

A retrospective review of consecutive mechanical thrombectomy cases was conducted at two high-volume regional stroke centers under institutional review board approved protocol (Sutter Health IRB number 2199694-1). All cases performed between January 2019 and December 2025 were reviewed. When available, stored fluoroscopy loops and stored monitor images were reviewed to assess aspiration catheter tip behavior during inner device removal for each thrombectomy pass. Images were considered adequate if a fluoroscopy loop included both the delivery of the inner device followed by the aspiration catheter to the angiographic target, as well the final position after removal of the inner device. Additionally, so operators would also save single-shot images or additional fluoroscopy loops after connecting to vacuum suction and starting the thrombectomy pass. These data were also reviewed when available. Passive advancement was defined as antegrade movement of the aspiration catheter tip during or immediately following inner device removal. This was distinguished from active operator-directed catheter advancement by reviewing the fluoroscopic sequence and confirming the temporal relationship between inner device movement and catheter tip displacement, as well as review of the procedural reports. It is the standard practice of all providers to not actively advance the aspiration catheter over the inner device, with exceptions of aspiration performed with no inner device, which is accounted for in the analysis. Data were reviewed by two reviewers with extensive neurointerventional experience with consensus made on cases of disagreement.

For passes in which passive advancement was confirmed or for which a post-inner-device-removal image was available, the final aspiration catheter tip position was recorded as located at an arterial branch point or not based on the subsequent angiogram delineating anatomy beyond the occlusion. Branch point positioning was defined as the catheter tip positioned at an identifiable arterial branch point on the subsequent angiographic run following the successful thrombectomy pass. Branch-point assessment was performed for all passes, irrespective of whether sufficient images were present to separately assess if passive advancement had occurred. For instance, if an image was not saved prior to inner device withdrawal but an image was saved demonstrating final aspiration catheter tip position, branch-point assessment was performed but passive advancement could not be assessed. Representative fluoroscopic images illustrating the assessment approach are shown in Figure 1. Given this assessment of branch points on subsequent images, reviewers were not blinded to recanalization outcomes.

**Figure 1.**
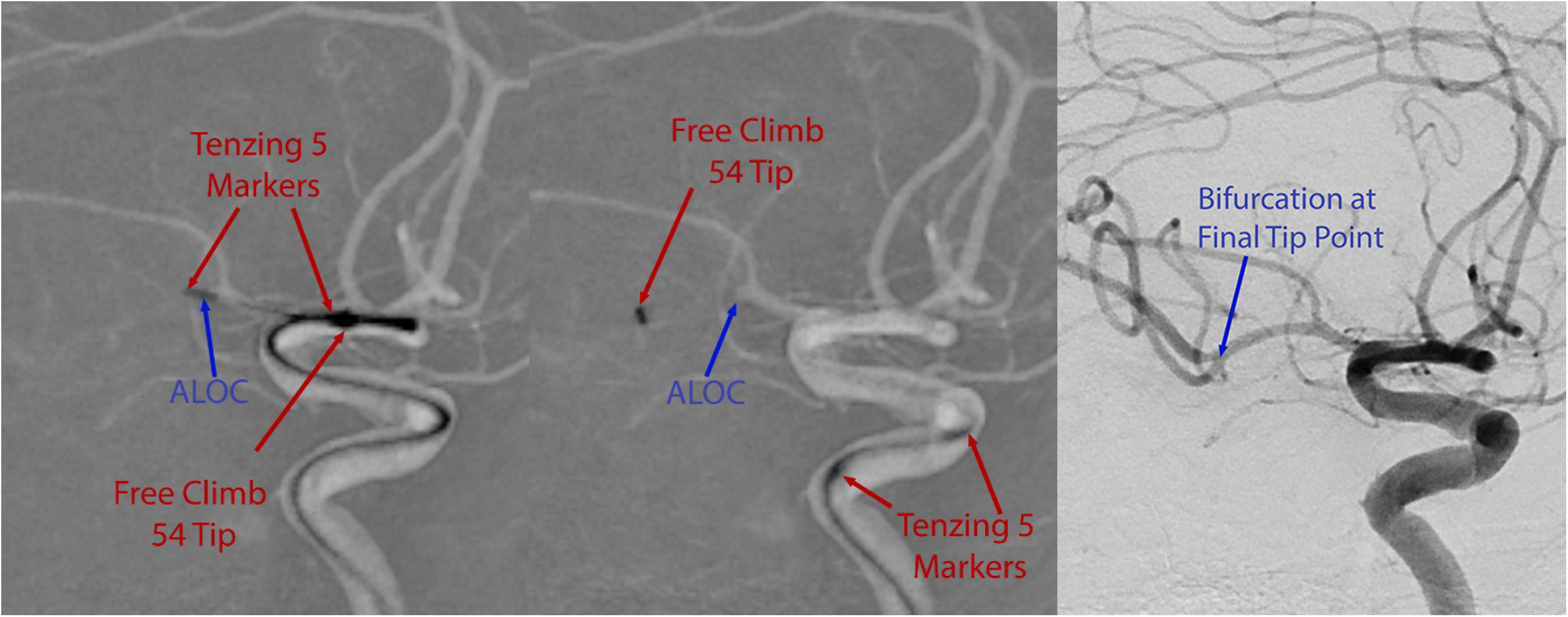
Representative fluoroscopic images demonstrating passive advancement assessment methodology. Left: Pre-withdrawal image with the aspiration catheter (FreeClimb 54 Tip) at the level of the angiographic limit of contrast (ALOC) and the inner delivery device (Tenzing 5 markers) positioned distally past the clot. Middle: Post-withdrawal image showing the aspiration catheter tip has advanced distally beyond the original ALOC — confirming passive advancement — while the inner device has been retracted proximally. Passive advancement was defined as antegrade catheter movement during or immediately following inner device removal, confirmed by comparing catheter tip position relative to the ALOC across sequential images. Right: Post-treatment angiogram demonstrates an arterial branch point at the final aspiration catheter tip location.

Primary outcomes were eTICI score, eTICI ≥2b-3, and eTICI ≥2c-3. FPE (eTICI ≥2c-3) and mFPE (eTICI ≥2b-3) were analyzed on a per-vessel (first-pass) basis. Secondary outcomes were secondary occlusion, which could be either a new thromboembolic occlusion in a previously uninvolved territory or residual thrombus from the originally involved thrombus that was not completely removed, and procedural complications. Fisher’s exact test was used for univariable categorical analysis. Per-pass and per-vessel analyses were performed separately to account for the nested data structure of multiple passes per patient and to distinguish technique-specific effects from outcome-level effects. Mixed-effects multivariable logistic regression was performed to identify independent associations while controlling for covariates including technique, access route, aspiration catheter and inner device type, occlusion location, intravenous thrombolytic status, tandem disease, and intrinsic intracranial atherosclerotic disease (ICAD). Multivariable models are summarized in Supplementary Table 1. Statistical significance was set at p=0.05; all tests were two-tailed.

## RESULTS

The overall cohort comprised 1,027 occlusions in 995 consecutive patients and 1,824 thrombectomy passes. Of 1,824 total passes, 733 (40.2%) had imaging adequate for passive advancement assessment, including 429 first passes. Branch position data were available for 1,448 passes (79.4%) in the full cohort. Passive advancement was observed in 525/733 assessed passes (71.6%) and passive advancement to an arterial branch in 306/733 (41.7%). Among the 429 first passes with imaging permitting passive advancement assessment, passive advancement was observed in 336/429 (78.3%), reaching an arterial branch point in 218/429 (50.8%). Patient demographics are summarized in Table 1.

**Table 1.** Patient Demographics.

| Characteristic | Overall Cohort (N=995 patients; 1,027 occlusions) |
| --- | --- |
| Age, mean ± SD | 70.2 ± 15.6 years |
| Female sex | 540/995 (54.3%) |
| IV alteplase | 269/1,019 (26.4%) |
| Left hemisphere occlusion | 582/1,027 (56.7%) |
| Right hemisphere occlusion | 400/1,027 (38.9%) |
| Tandem disease | 174/1,027 (16.9%) |
| Intrinsic ICAD | 102/1,027 (9.9%) |
| Occlusion location |  |
| ICA | 151 (14.7%) |
| M1 | 390 (38.0%) |
| M2 | 178 (17.3%) |
| M3 | 184 (17.9%) |
| M4/distal MCA | 5 (0.5%) |
| ACA (A2–A4) | 29 (2.8%) |
| Basilar artery | 45 (4.4%) |
| PCA (P1–P3) | 37 (3.6%) |
| Vertebral (V4) | 8 (0.8%) |

ICAD, intracranial atherosclerotic disease. †Age was available for 260/1,027 vessel-level records (mean reflects patients with non-zero age entry); sex from patient-level tallies (N=995). NIHSS was not collected. Occlusion location percentages based on 1,027 occlusions; all other proportions use the denominator specified in each row.

On per-pass analysis (n=733 passes with passive advancement data), passive advancement was associated with higher eTICI ≥2b-3 compared to passes without passive advancement (71.0% vs 19.7%, p<0.001); this was also found for eTICI ≥2c-3 (57.5% vs 10.1%, p<0.001). When passive advancement reached an arterial branch point, these associations were amplified with eTICI ≥2b-3 in 90.5% vs 32.1% and eTICI ≥2c-3 in 79.4% vs 18.7% (both p<0.001). These data are summarized in Table 2.

**Table 2.** Passive Advancement Outcomes — Per-Pass Analysis.

| Outcome | PA Present | PA Absent | PA to Branch | No PA to Branch | p-value |
| --- | --- | --- | --- | --- | --- |
| Per-Pass Analysis (n=733) |  |  |  |  |  |
| n | 525 | 208 | 306 | 427 |  |
| eTICI ≥ 2b-3 | 71.0% | 19.7% | 90.5% | 32.1% | <0.001 (all) |
| eTICI ≥ 2c-3 | 57.5% | 10.1% | 79.4% | 18.7% | <0.001 (all) |

PA, passive advancement; FPE, first-pass effect (eTICI ≥ 2c-3 on first pass); mFPE, modified first-pass effect (eTICI ≥ 2b-3 on first pass). For PA columns: PA Present vs PA Absent. For branch columns: PA to Arterial Branch vs No PA to Arterial Branch (includes passes with and without PA that did not reach a branch). All p-values from Fisher's exact test.

On first-pass analysis (n=429 first passes with passive advancement assessment imaging available), mFPE was 74.1% with versus 10.8% without passive advancement (p<0.001), and FPE was 63.4% vs 7.5% (p<0.001). When passive advancement reached an arterial branch point, mFPE was 95.0% versus 24.6% without advancement to a branch (p<0.001), and FPE was 87.2% vs 14.2% (p<0.001). These findings are also summarized in Table 3.

**Table 3.** Passive Advancement Outcomes — First-Pass Analysis.

| Outcome | PA Present | PA Absent | PA to Branch | No PA to Branch | p-value |
| --- | --- | --- | --- | --- | --- |
| First-Pass Analysis (n=429) |  |  |  |  |  |
| n | 336 | 93 | 218 | 211 |  |
| mFPE (eTICI $\geq 2b-3$) | 74.1% | 10.8% | 95.0% | 24.6% | <0.001 (all) |
| FPE (eTICI $\geq 2c-3$) | 63.4% | 7.5% | 87.2% | 14.2% | <0.001 (all) |

*PA, passive advancement; FPE, first-pass effect (eTICI $\geq 2c-3$ on first pass); mFPE, modified first-pass effect (eTICI $\geq 2b-3$ on first pass). For PA columns: PA Present vs PA Absent. For branch columns: PA to Arterial Branch vs No PA to Arterial Branch (includes passes with and without PA that did not reach a branch). All p-values from Fisher's exact test.*

On per-pass analysis, passive advancement to an arterial branch point was inversely associated with secondary occlusion (13.3% vs 31.3% without advancement to a branch point, p<0.001). This association suggests that antegrade catheter movement to a branch point by passive advancement during and/or after delivery device withdrawal may promote more complete clot ingestion, thus reducing fragmentation or distal migration of the clot, which corroborates the above described first-pass results. Furthermore, in per-pass analysis of all 1,448 passes where arterial branch points were documented— regardless of whether assessment of passive advancement was possible—final aspiration catheter position at an arterial branch point was associated with eTICI ≥2b-3 (85.1% vs 36.1%), eTICI ≥2c-3 (74.8% vs 25.9%), and lower secondary occlusion (15.5% vs 29.8%; all p<0.001).

Multivariable models are summarized in the supplementary materials table. On per-pass multivariable analysis, passive advancement was independently associated with higher eTICI score (p<0.001), eTICI ≥2b-3 (p<0.001), and eTICI ≥2c-3 (p<0.001). Final catheter position at an arterial branch point was independently protective against secondary occlusion per pass (p=0.042). On per-vessel multivariable analysis, final catheter position at an arterial branch point was the dominant independent predictor of FPE (p<0.001) and mFPE (p<0.001) and was independently inversely associated with secondary occlusion (p<0.001), any complication (p=0.046), and intracranial complication (p=0.038). Passive advancement to a branch point remained independently protective against secondary occlusion in per-vessel analyses (p=0.042). Multivariable analysis results are summarized in Supplemental Tables 2 and 3.

Device-specific analysis demonstrated no independent association between individual aspiration catheter or inner delivery device models and recanalization or safety outcomes in either per-pass or per-vessel multivariable analysis. However, the use of manufacturer-paired aspiration catheters and inner delivery devices—defined as both devices sourced from the same manufacturer—was independently associated with reduced secondary occlusion on per-vessel multivariable analysis (β=−0.052, 95% CI: −0.094 to −0.010; p=0.016). Paired device use was not independently associated with any outcome on per-pass multivariable analysis. A summary of device utilization across the cohort is provided in Supplemental Table 4, and the effect of manufacturer-paired device use across all multivariable models is presented in Supplemental Table 5.

Inner delivery device type—classified as guidewire, standard catheter, or tapered delivery device—affected passive advancement rates. Passive advancement occurred in 54.1% of passes using a guidewire, 60.7% with a microcatheter, and 79.6% with a tapered delivery device (p<0.001). Passive advancement to an arterial branch point occurred in 24.6%, 28.3%, and 50.3% of passes, respectively (p<0.001). Final catheter tip position at an arterial branch was achieved in 28.7% of passes using a guidewire, 16.2% with a standard microcatheter, and 50.2% with a tapered delivery device (p<0.001; N=1,440). On per-pass multivariable analysis, tapered delivery device use was independently associated with higher eTICI ≥2c-3 compared to guidewires and standard microcatheters (β=0.082, 95% CI: 0.011 to 0.154; p=0.024), with a borderline association with eTICI ≥2b-3 (β=0.072, p=0.073); no independent association was identified between inner delivery device type and secondary occlusion or any complication. Per-pass recanalization and secondary occlusion outcomes by individual inner devices are provided in Supplemental Table 6.

To assess for potential selection bias introduced by the requirement for adequate fluoroscopic imaging, baseline and outcome characteristics were compared between the 733 passes with adequate imaging and the 1,091 passes without adequate imaging (Supplemental Table 7). Passes with adequate imaging had higher rates of radial access (11.2% vs. 3.4%), lytic therapy (29.1% vs. 24.5%), and final catheter position at an arterial branch (43.7% vs. 25.7%), and lower rates of tandem disease (12.0% vs. 21.8%) and inner device crossing past the clot (35.9% vs. 58.3%). Fewer ICA occlusions occurred in the assessed group (15.8% vs. 21.9%). These differences likely reflect temporal and site-based variation in imaging storage practices rather than systematic case selection, particularly given hardware upgrades through the study period that enhanced fluoroscopic loop storage capability. Non-assessed passes were disproportionately concentrated in earlier time periods and device generations when fluoroscopic clip storage was less feasible and inconsistent. The primary outcome measures were largely comparable between groups. eTICI ≥2c-3 did not differ significantly (44.1% vs. 41.6%, p=0.299), and secondary occlusion (24.8% vs. 25.7%, p=0.688) and complication rates (1.8% vs. 2.0%, p=0.711) were similar. A difference in eTICI ≥2b-3 was observed (56.5% vs. 50.0%, p=0.006).

## DISCUSSION

This study provides strong evidence that passive advancement of an aspiration catheter is a prevalent, fluoroscopically identifiable phenomenon that is independently associated with more favorable recanalization results. Advancement of the aspiration catheter to an arterial branch point achieved FPE (eTICI ≥2c-3) in 87.2% of first passes—among the highest single-pass reperfusion rates reported in any aspiration thrombectomy study^13^. Final aspiration catheter position at an arterial branch was the dominant independent predictor of most measured outcomes on per-vessel multivariable analysis.

To our knowledge, passive advancement of the aspiration catheter upon inner delivery device withdrawal has not been previously named or systematically evaluated as a technically advantageous phenomenon. While analogous catheter advancement during inner device withdrawal in other endovascular procedures is widely recognized, this is the first study to demonstrate the clinical significance of passive advancement in the context of aspiration thrombectomy, and in relation to arterial branch points, on angiographic outcomes. The traditional operator instinct to resist unintended antegrade catheter movement has historically been trained behavior, motivated by caution about uncontrolled advancement in the fragile intracranial circulation. Our data suggest that this instinct is counterproductive in the aspiration thrombectomy context; passive advancement should be actively identified and embraced when occurring safely with the use of modern delivery devices; it was not associated with higher complication rates in aspiration thrombectomy.

These observations extend prior evidence that distal catheter positioning improves recanalization. In combined stent retriever-aspiration thrombectomy, distal access catheter tip positioning adjacent to the thrombus achieves superior complete reperfusion versus proximal positioning (66.7% vs 42.1%, p=0.025).^11^ Higher intracranial positioning of guide catheters has similarly been associated with improved first-pass aspiration success.^12^ However, the magnitude of the association observed here—with first-pass eTICI ≥2b-3 of 95.0% at arterial branch points—substantially exceeds the effect sizes in these prior reports and represents a new maximum for single-factor recanalization prediction in the aspiration thrombectomy literature.

The stronger recanalization associations when advancement reaches an arterial branch (mFPE 95.0% and FPE 87.2% on first-pass analysis versus 74.1% and 63.4% with passive advancement not to an arterial branch point, respectively) suggest that the aspiration catheter reaching an arterial branch point positioning creates favorable conditions for clot extraction. Advancement to a branch point may be an indicator of an aspiration catheter advancing unimpeded through the lumen of an artery. Similarly, a thromboembolus is also likely to lodge at a bifurcation, so advancement to a branch point increases the likelihood that the catheter position corresponds to the true site of occlusion. Once the aspiration catheter is delivered to the proximal aspect of a thrombus, the catheter likely engulfs the thrombus into its tip as it advances passively. Additionally, as vacuum is created at the distal tip of the catheter with removal of the inner delivery device, this further potentiates recanalization by starting aspiration immediately as the catheter reaches the occlusion. Moreover, the advancement of aspiration catheters after delivery device removal to an arterial bifurcation point and subsequent cessation of catheter movement without causing damage likely reflects increased catheter tip softness exhibited by numerous devices from multiple manufacturers utilized in this series. The angiographic limit of contrast on digital subtraction angiography may not actually represent the site of occlusion if a column of stagnant contrast exists just upstream to the occlusion. Catheter advancement to a branch point may also indicate that the catheter is advancing to the true location of the thrombus. The branch point geometry may constrain the clot at this site, with thrombus size preventing it from lodging into a more distal branch with a smaller lumen, in effect pinning the thrombus in place so the aspiration catheter can engulf and aspirate it. Passive advancement is a phenomenon that should be observed but not actively sought out. Similarly, the branch position phenomenon should be considered a post-facto marker of effective passive advancement and its attendant aspiration benefits. It should be emphasized that active force applied by the operator to the aspiration catheter to induce advancement could be dangerous and lead to serious complications.

The independent association between passive advancement to a branch and reduced secondary occlusion (13.3% vs 31.3%, p<0.001; independently protective in per-pass multivariable analysis, p=0.042) also supports the mechanistic arguments described above. It follows that a catheter tip that passively advances to engulf a thrombus lodged at a bifurcation is more likely to ingest the clot intact. This mechanical pre-containment of a clot in the catheter tip may reduce the tendency of the clot to fragment, dislodge, or migrate downstream beyond the bifurcation. Additionally, aspiration catheters adequately sized to the vessel lumen are more likely to temporarily arrest flow, which may also limit downstream migration if fragmentation occurs.

The protective effect of final catheter position at an arterial branch against any complication (p=0.046) and intracranial complication (p=0.038) on per-vessel analysis is noteworthy. Catheter antegrade movement due to passive advancement within the intracranial vasculature, without a leading inner device, might be expected to increase, rather than reduce, procedural risk. The most likely explanation of this safety finding is that aspiration catheters reaching an arterial branch point after passive advancement arrive there transiting through the arterial lumen without sufficient friction to arrest antegrade motion, until the change in vessel caliber present at an arterial branching point. Importantly, in this study the passive advancement phenomenon did not cause an increase in angiographically visible signs of clinically relevant endothelial damage, such as dissection or perforation, compared to cases in which passive advancement was not observed.

This study has several limitations to address. Passive advancement could be assessed in 733 of 1,824 passes (40.2%), reflecting inconsistent fluoroscopy clip storing; cases with available imaging may differ systematically from those without and this may have introduced bias. However, largely similar patient, procedural, and outcomes variables between passes with and without adequate imaging to assess passive advancement suggests this is unlikely to represent meaningful selection bias. The definition of passive advancement required author review and is subject to inter-rater variability; no formal reliability testing nor core lab adjudication was performed. Lack of blinding to recanalization results could also introduce bias. The retrospective design precludes causal inference. The cohort spans 7 years across multiple aspiration catheter systems, operators, and technique regimes. This introduces heterogeneity in how passive advancement was recognized and recorded, and these results may not be generalized to other aspiration catheter platforms or mechanical thrombectomy techniques.

## CONCLUSIONS

Passive advancement of the aspiration catheter upon inner delivery device withdrawal is a prevalent physical phenomenon and independently predicts favorable recanalization following aspiration thrombectomy. Both passive advancement and passive advancement of the aspiration catheter to an arterial branch point are associated with high FPE and mFPE and were found to be independent predictors of recanalization success, secondary occlusion prevention, and reduced complications. These results suggest that passive advancement after delivery device removal, and selection of devices and techniques that produce it, is a desirable phenomenon and should be incorporated into aspiration thrombectomy technical considerations. Further study is warranted.

## Data Availability

The data supporting this study are available from the corresponding author upon reasonable request, subject to institutional IRB approval and applicable privacy regulations.

## COMPETING INTERESTS

MDA: Consulting and equity, Certus Critical Care; Consulting and equity, Route 92 Medical; Consulting, Stryker; Consulting, Medtronic; Consulting, J&J; Consulting and equity, Piraeus Medical; Equity, Galaxy Therapeutics. WTK: Consultant fees, Stryker Neurovascular; Consultant fees, Travel reimbursements, and Stock Options, Route 92 Medical. FS: Consultant fees, Stryker Neurovascular and Route 92 Medical; Honoraria for lectures, Stryker Neurovascular; Travel, Medtronic; Research Grants: Microvention, Stryker. Stock or Stock Options: Route 92 Medical. BV, TCC, RSK: None.

## FUNDING

Not applicable. There are no funders to report for this submission.

## Notes

### Author Declarations

Sutter Health Institutional Review Board

